# A Hypothalamus centered Pathogenesis of Heat Stroke Deaths- A Postmortem-Based Human Study

**DOI:** 10.1101/2025.03.10.25323319

**Authors:** Ashok k Rastogi, Ashutosh Kumar, Shreekant Bharti, Sanjeev K Paikra, Ashok K Datusalia, Amit M Patil, Rajesh Kumar

**Affiliations:** Department of Forensic Medicine & Toxicology, All India Institute of Medical Science, Patna (Bihar), India; Department of Anatomy, All India Institute of Medical Sciences, Patna; Department of Pathology, All India Institute of Medical Science, Patna (Bihar), India; National Institute of Pharmaceutical Education and Research (NIPER) Raebareli

**Keywords:** Heat stroke, Hypothalamus, Death, Summer, Hemorrhage

## Abstract

**Background:** Heat stroke deaths are increasing during the summer in tropical countries with hot climates. The pathogenesis mechanisms leading to death in heat stroke are not well established. In addition, postmortem-based human studies in heat stroke cases are scarce in the literature, which makes the forensic determination of the cause of death extremely difficult in autopsied cases.

**Materials and Methods:** The four deceased cases (three male and one female, age range: 40-60 years) with a history of heat exposure were received in the mortuary for forensic determination of the cause of death. The autopsy of the cases was performed using standard protocol and gross observations were made. Further, the visceral tissue samples were taken for histological staining and microscopic examination.

**Results:** In gross examination, hemorrhage, and acute vascular and necrotic lesions were observed in the preoptic area of the anterior hypothalamus bilaterally. Multiple hemorrhagic foci were also observed over the external surfaces of the heart and lungs.

In microscopic examination, hemorrhagic foci were observed in the hypothalamus, cerebellum, pons, and medulla. In addition, acute vascular lesions and tissue necrosis were observed in the heart, lungs, kidney, and liver.

**Conclusions:** Our observations indicate that the anterior hypothalamus may have a commanding role in the pathogenesis of heat stroke. Acute hemorrhage in the preoptic area of the anterior hypothalamus associated with microvascular injury and acute necrotic lesions of key organs involved in the hemodynamic circulation and temperature control, primarily the heart, lung, kidney, and liver should be considered in the forensic determination of the cause of death in such cases.

## Introduction

Scorching heat waves during the summer are becoming increasingly common in hot-climate tropical countries. A score of people lose their lives every year in these regions due to exposure to excessive heat. When the core body temperature exceeds 40 degrees Celsius, it leads to hot and dry skin and dysfunction of the central nervous system and vital organs involved in hemodynamic circulation and temperature control, leading to heat stroke^1^ Elevated core body temperature can lead to significant mortality in the absence of required care. In heat stroke, the hypothalamus supposedly drives the central nervous system dysfunction, which is further complicated by multiorgan dysfunction^2,3^ Notably, the preoptic area of the anterior hypothalamus is a key center for heat regulation.^4^The extensive damage to preoptic nuclei accompanied by the multi-organ dysfunction is well documented in the animal model studies of heat stroke. However, the literature on such pathological changes in human cases is scarce. A standard autopsy protocol to confirm the death due to heat stroke is also missing in the literature. In this study, we aim to establish the tissue injury pattern and key pathological mechanisms leading to death in heat stroke cases.

## Materials and Methods

History taking, autopsy examinations, and preservations of samples:

### History of cases

The four deceased cases (three male and one female, age range: 40-60 years) with a history of heat exposure were received in the mortuary for forensic determination of the cause of death.

On the day of heat exposure, the maximum temperature in the area reached 48 degrees Celsius, while the minimum temperature was 33 degrees Celsius, as per the city’s meteorological service records. Humidity peaked at 95% and dropped to a low of 65%. The relatives and police personnel gave the history to provide information regarding exposure to heat and death. The average environmental heat exposure for each case lasted 5-8 hours.

The deceased were externally examined for any hemorrhage, discoloration of the skin, changes in the eye, or any other abnormality. An incision was given from the chin to the pubis symphysis. The chest, abdominal, and cranial cavities were opened. All major organs, such as the lungs, liver, spleen, kidney, and brain, were examined, and observations were noted down. Based on the gross observations, the tissue samples from the internal organs were harvested and preserved in 10% buffered formalin for further microscopic examination.

### Gross and microscopic examinations

During the autopsy examination, macroscopic changes in internal organs were observed. Microscopic observation was done after the Hematoxylin & Eosin (H&E) staining, and observations were made under a bright-field light microscope. The inferences were drawn from observations of gross and histological examinations.

Statistical evaluations: The percentage of the observations was calculated among the examined cases.

### Ethical statement

Ethical clearance was obtained wide letter, no AIIMS/Pat/IEC/2023/1162 from the ethical committee from the All-India Institute of Medical Sciences Patna

## Results

### Gross observations (Figure 1, Tables 1-3)

Nail bed cyanosis and congestion were present in all (100%) cases; however, no clear hemorrhagic spots were observed in the skin. Hemorrhage over the preoptic area of the hypothalamus was observed in two out of three cases (66.67%). The organs were found congested. The vessels were dilated and thrombosed. Multiple hemorrhagic foci were observed over the external surface of the heart and lungs, and white matter of the brain. In the preoptic area of the brain, a small hemorrhagic patch measuring 1.5×0.1 cm was observed. Heart vessels were found dilated. The endocardial hemorrhage was noted in one case.

**Table 1.**
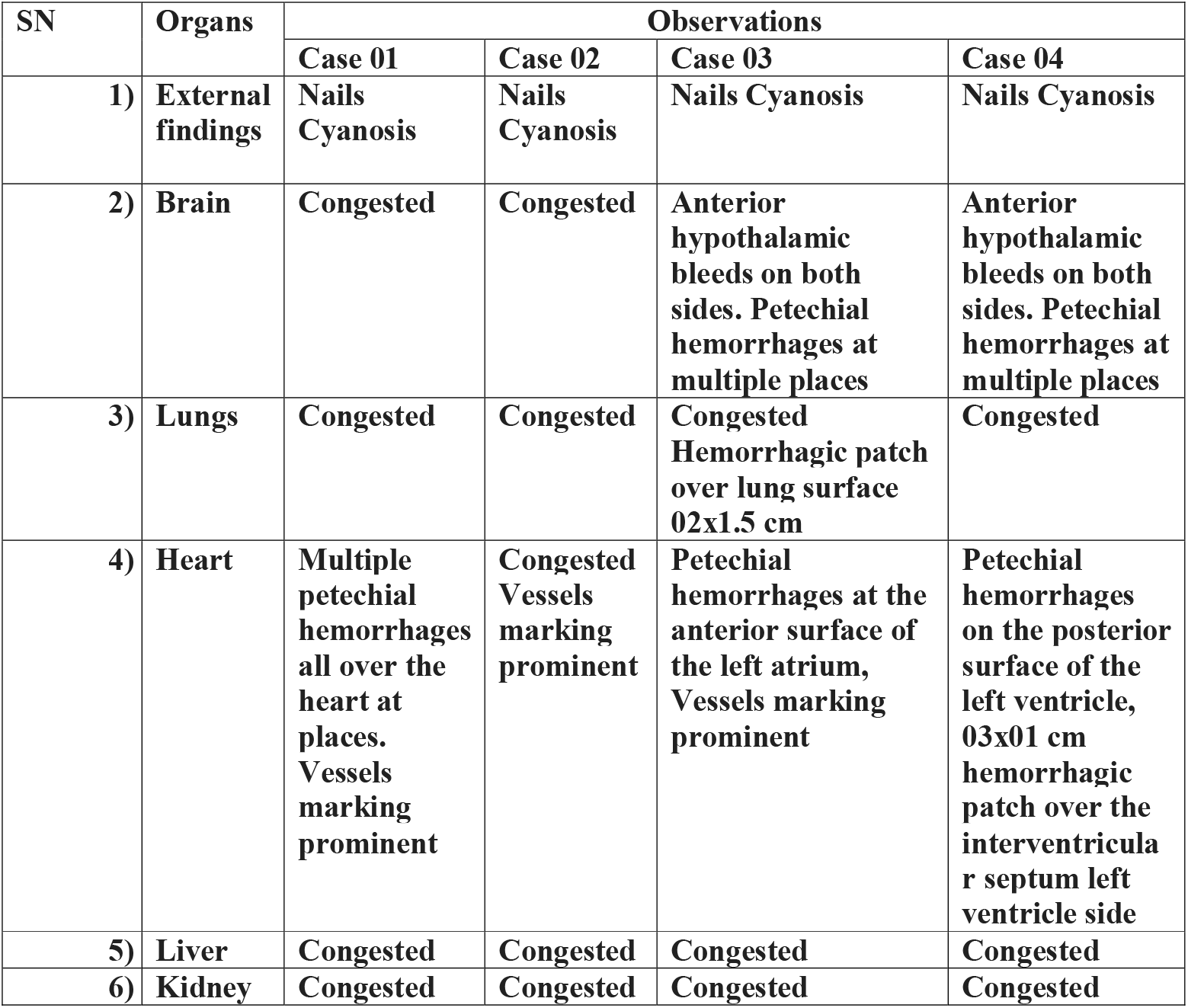
Observations during autopsy examination.

| SN | Organs | Observations |  |  |  |
| --- | --- | --- | --- | --- | --- |
|  |  | Case 01 | Case 02 | Case 03 | Case 04 |
| 1) | External findings | Nails Cyanosis | Nails Cyanosis | Nails Cyanosis | Nails Cyanosis |
| 2) | Brain | Congested | Congested | Anterior hypothalamic bleeds on both sides. Petechial hemorrhages at multiple places | Anterior hypothalamic bleeds on both sides. Petechial hemorrhages at multiple places |
| 3) | Lungs | Congested | Congested | Congested Hemorrhagic patch over lung surface 02x1.5 cm | Congested |
| 4) | Heart | Multiple petechial hemorrhages all over the heart at places. Vessels marking prominent | Congested Vessels marking prominent | Petechial hemorrhages at the anterior surface of the left atrium, Vessels marking prominent | Petechial hemorrhages on the posterior surface of the left ventricle, 03x01 cm hemorrhagic patch over the interventricular septum left ventricle side |
| 5) | Liver | Congested | Congested | Congested | Congested |
| 6) | Kidney | Congested | Congested | Congested | Congested |

**Table 2.**
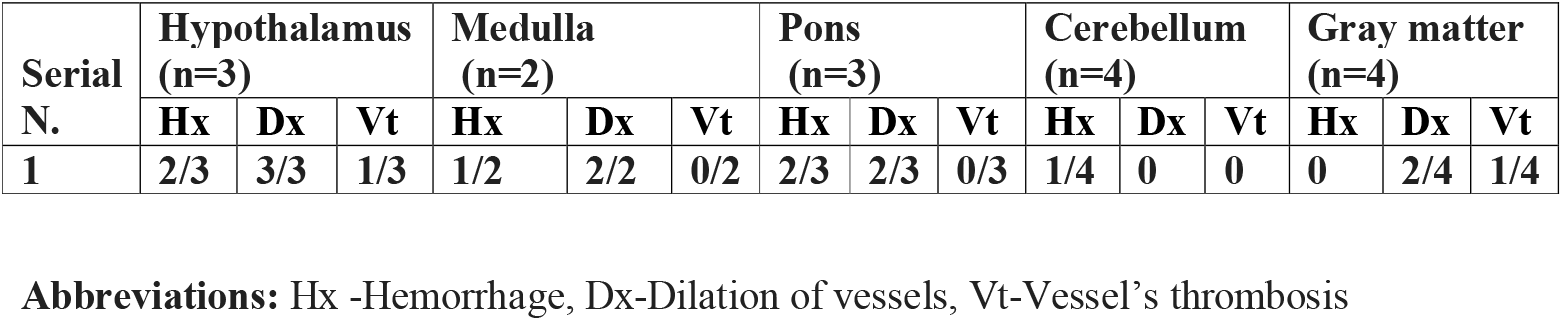
Observations in the brain.

**Table 3.**
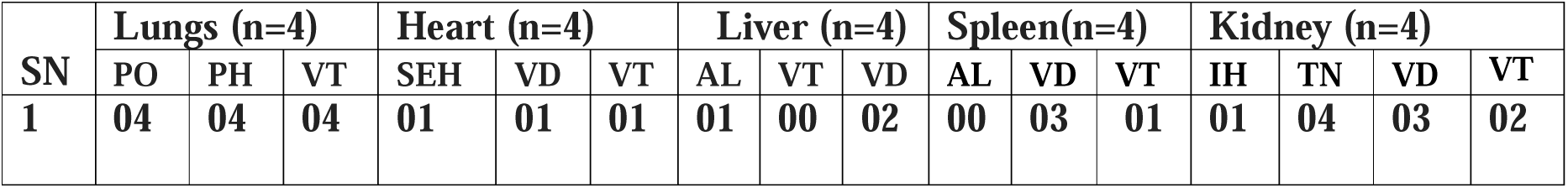
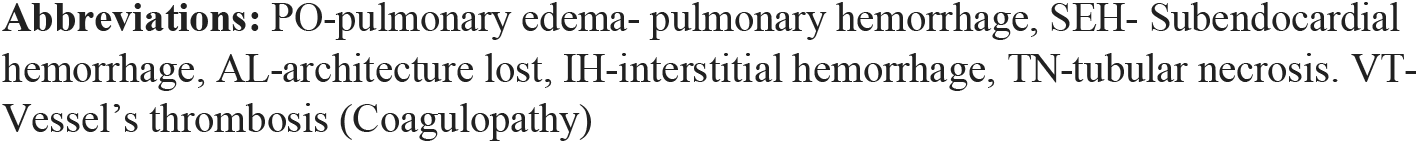
Observation in the other organs.

| SN | Lungs (n=4) |  |  | Heart (n=4) |  |  | Liver (n=4) |  |  | Spleen(n=4) |  |  | Kidney (n=4) |  |  |  |
| --- | --- | --- | --- | --- | --- | --- | --- | --- | --- | --- | --- | --- | --- | --- | --- | --- |
|  | PO | PH | VT | SEH | VD | VT | AL | VT | VD | AL | VD | VT | IH | TN | VD | VT |
| 1 | 04 | 04 | 04 | 01 | 01 | 01 | 01 | 00 | 02 | 00 | 03 | 01 | 01 | 04 | 03 | 02 |

**Abbreviations:** PO-pulmonary edema- pulmonary hemorrhage, SEH- Subendocardial hemorrhage, AL-architecture lost, IH-interstitial hemorrhage, TN-tubular necrosis. VT- Vessel's thrombosis (Coagulopathy)

**Figure 1.**
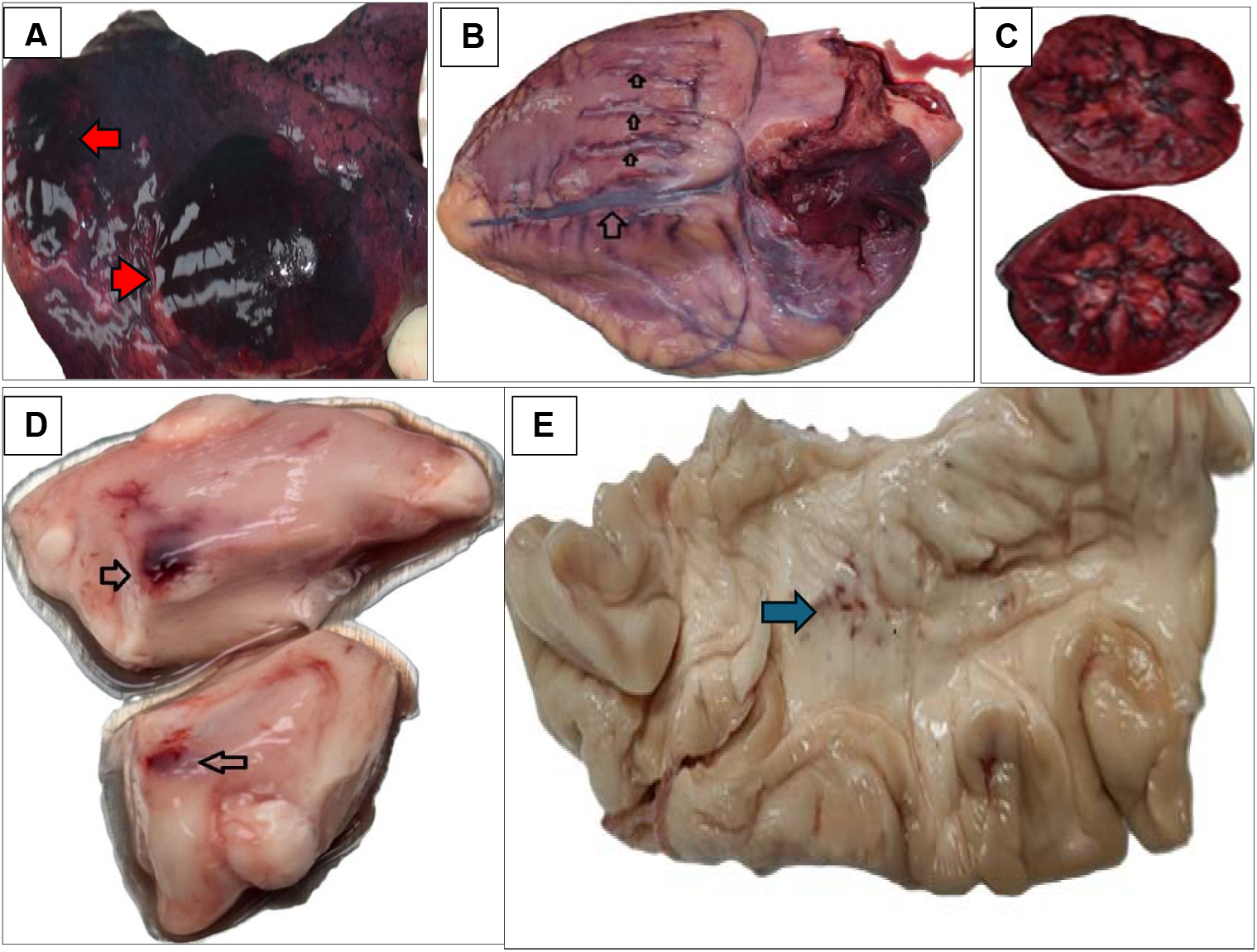
Gross autopsy observations in internal organs, A-Hemorrhagic patch over the surface of the lung (Red Arrow), B-Over the heart surface vessels dilated and prominent (hollow black arrow). C-Highly congested kidneys. D- Both sides of the anterior hypothalamus hemorrhage and congestion (Hollow black arrow). E-Petechial hemorrhages are seen in the brain’s white matter (Blue arrow).

### Microscopic observations (Figures 2 and 3, Tables 2 and 3)

Microscopic hemorrhagic foci were observed in the hypothalamus, cerebellum, pons, and medulla (Fig. 2: a, b, c, and d). Endocardial hemorrhage and myofibril necrosis were noted (Fig.3 c and d). In renal, Acute tubular necrosis with collecting duct necrosis was noted (Fig.3 a and b). The vessels were dilated and thrombosed in the lung tissue (Fig. 3e). The acute central necrosis with loss of architecture was noted in the liver (Fig.3f).

**Figure 2.**
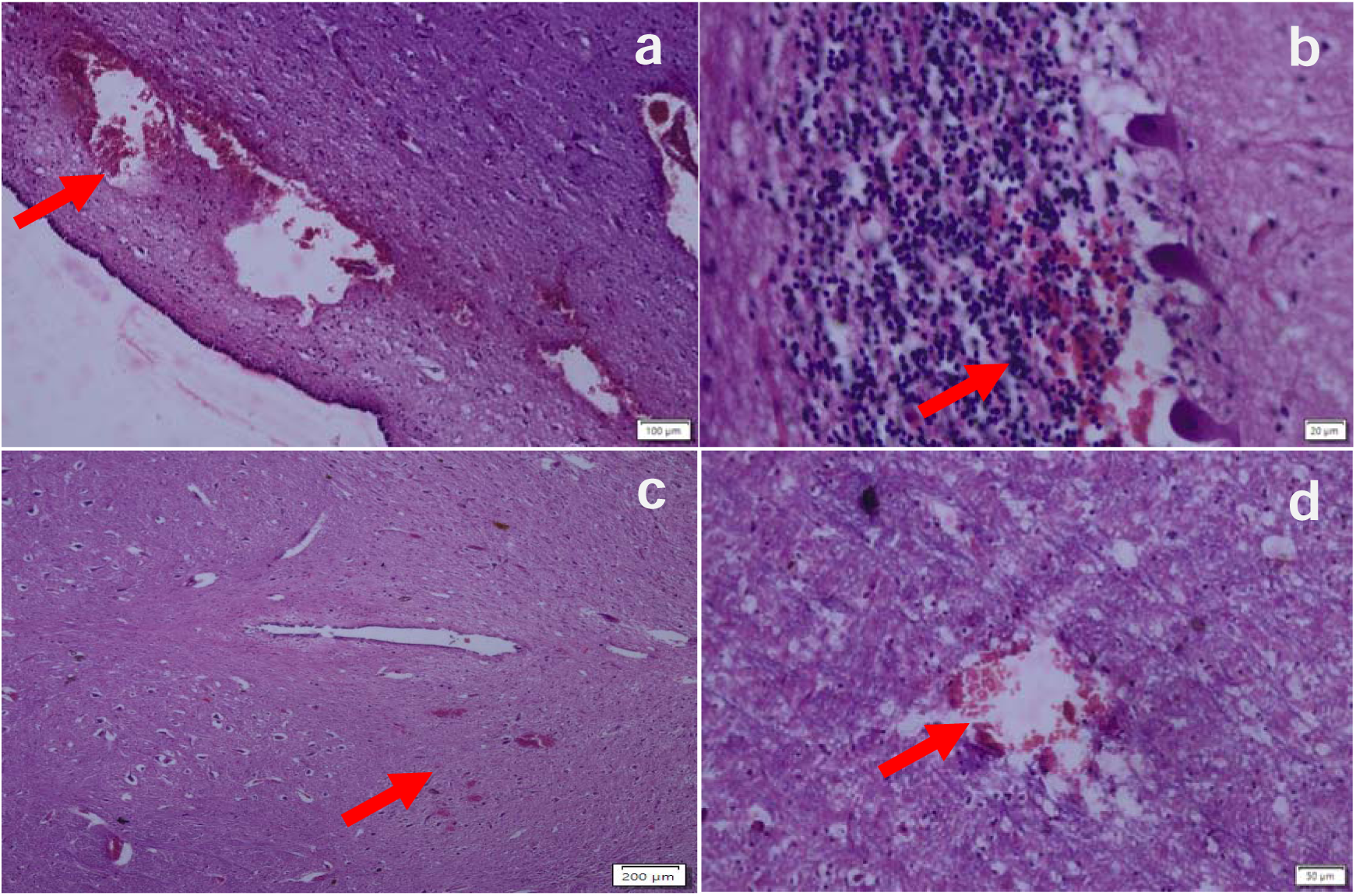
(a, b, c & d)-Showing hemorrhage indicated by red arrow in Hypothalamus, cerebellum, pons and medulla respectively.

**Figure 3.**
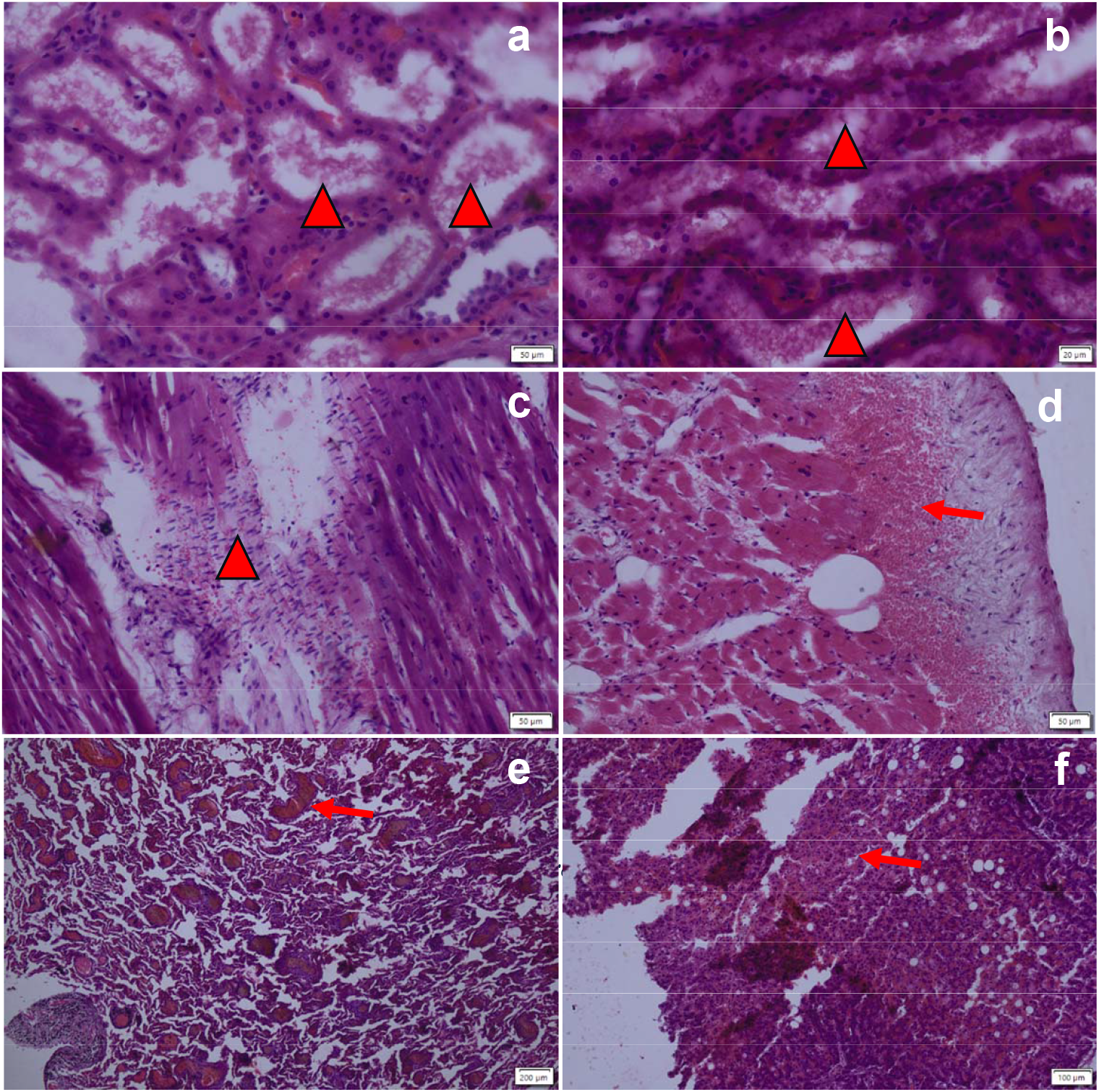
Microscopic observations in internal organs, a-kidney acute tubular necrosis (red arrowhead), b-Collecting duct necrosis (Red arrowhead), c-Cardiac muscle necrosis with neutrophilic infiltration (Red arrowhead), d-Endocardial hemorrhage (Red arrow), e-Lung vascular thrombosis and vascular hemorrhage (red arrow), f-Liver sinusoidal necrosis lost architecture (Red arrow).

## Discussion

Rising incidences of heat stroke deaths have posed a challenge for forensic experts to confidently establish the cause of death from the autopsy findings in such cases. Our study reveals a unique tissue injury pattern in postmortem examination of heat stroke cases, which may help in understanding its pathogenesis and establishing the cause of death. We observed macroscopic as well as microscopic evidence of acute vascular and necrotic lesions in the anterior hypothalamus, particularly in the preoptic area. Hemorrhagic and necrotic spots were also noted in the other brain regions, such as pons, medulla, and cerebellum, indicating the involvement of the pathogenesis of heat stroke.^5^ Other than the brain, microvascular injuries and tissue necrosis were noted in the vital organs involved in the control of hemodynamic circulation and body temperature, such as the lungs, heart, kidneys, and liver.

Although postmortem-based human studies are scarce, the microvascular injury in the hypothalamus is well documented in the animal model studies of heat stroke. ^2,6,7^ The anterior hypothalamus (preoptic area) bears the thermoregulatory center. In an attempt to lower the body temperature, the dilatation and burst rupture of the microvasculature of this region may occur in excessive heat exposure. ^8^

An increase in the surface and core body temperature following heat exposure activates the hypothalamic pituitary adrenal (HPA) pathway and, in turn, the autonomic nervous system (Fig. 4) through cytokine-mediated mechanisms. ^9,10^ In response, increased cardiac output, sweating, and a generalized dilatation of the microvasculature of the skin and other body organs ensues. ^9,10^ In response to the signals received from the peripherally and centrally located thermal receptors, a specific group of neurons in the preoptic area of the anterior hypothalamus releases antidiuretic hormone (arginine vasopressin). The antidiuretic hormone acts on the glomerular tubules of the kidney and directs a reduction in urine output, in turn, increases the total blood volume. ^9,10^ A generalized vascular dilatation and increased cardiac output also speed up the blood circulation across the body’s tissues and help dissipate the heat. ^9,10^ However, in case of excessive heat exposure, the hypothalamic control to bring down the body temperature may get overdriven and risk failure of the temperature-controlling mechanisms. A failure of the temperature control center may manifest as the rupture and bleeding in the micro-vessels in the anterior hypothalamic region and other organs involved in the hemodynamic circulation and temperature control,^9 10^ which has been evidenced in our study. Moreover, extreme heat exposure may also induce hemoconcentration, leading to intravascular coagulation. ^11^ Due to increased sweating, electrolyte balance may also ensue.^1^ Heat stroke-induced inflammatory reaction and vascular endothelial damage may further induce disseminated intravascular coagulation, resulting in microvascular injury and multi-organ failure. ^12–14^

**Figure 4:**
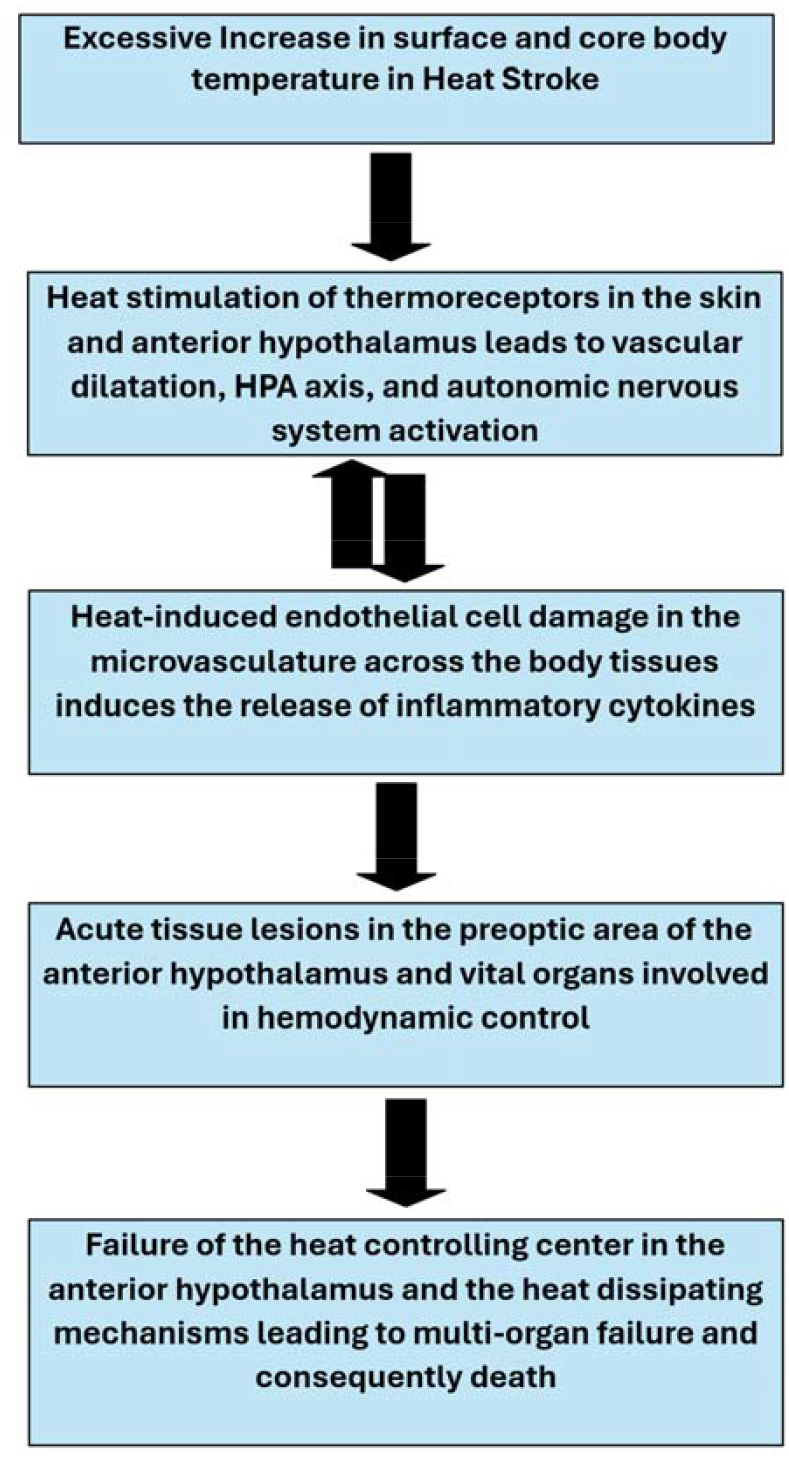
Schematic chart describing pathogenesis of heat stroke death.

The robust evidence of microvascular injuries and acute necrotic lesions localized to the heat-controlling region of the hypothalamus, accompanied by similar injuries in the internal organs, primarily in the heart, lung, kidney, and liver, is unraveled in our study. Our observations establish a unique tissue injury pattern in heat stroke deaths, which can be used as a signature for the forensic determination of the cause of death in autopsied cases. It also affirms a hypothalamus-mediated vascular overdrive of the organs involved in hemodynamic and temperature control as the key pathogenic mechanism leading to death in heat stroke cases.

## Conclusion

Our observations indicate anterior hypothalamus may have a commanding role in the pathogenesis of heat stroke. Acute hemorrhage and necrosis in the preoptic area of the hypothalamus associated with microvascular injury and acute necrotic lesions of key organs involved in the control of hemodynamic circulation and temperature control, primarily the heart, lung, kidney, and liver may be considered in the forensic determination of the cause of death in such cases.

## Limitations and further directions

Our conclusions are based on the observations made in limited cases of heat stroke. A larger sample size study may be necessary to validate the consistency of these findings. Further research is necessary to comprehend the inflammatory and molecular changes in the heat-regulating center of the anterior hypothalamus, other brain regions, and the internal organs involved in heat control, which will enhance our understanding of the pathogenesis of heat stroke deaths.

## Data Availability

all available in manuscript

## Acknowledgement

The authors are grateful to the kin of the deceased, who provided consent for collecting and reporting the clinical data.

## Funding agency

None

## Authors’ contributions

AKR and AK conceptualised the study. AKR and AMP carried out data collection. AKD and SKP performed laboratory work. AKR, SB, and AK conducted pathological validation and analysed the data. AKR and AK wrote the first draft of the manuscript. All authors reviewed and approved the final version of the manuscript

## References

1. Bouchama A and James P.K Nochel. Heat Stroke. N Engl J Med. 2002;346(25):1978–1988.

2. Chen S-H, Lin M-T, Chang C-P. Ischemic and Oxidative Damage to the Hypothalamus May Be Responsible for Heat Stroke. Curr Neuropharmacol. 2013;11(2):129–140. doi:10.2174/1570159x11311020001

3. Morgan L.O. Var. The hypothalmic nuclei heat stroke. Acta radiol. 1972;12(S 315):7–10. doi:10.1080/05678067209173960

4. Romanucci M, Della Salda L. Pathophysiology and pathological findings of heatstroke in dogs. Vet Med Res Reports. 2013;4:1–9. doi:10.2147/vmrr.s29978

5. Yoneda K, Hosomi S, Ito H, et al. How can heatstroke damage the brain? A mini review. Front Neurosci. 2024;18(October):1–8. doi:10.3389/fnins.2024.1437216

6. Chang CY, Chen JY, Chen SH, Cheng TJ, Lin MT, Hu ML. Therapeutic treatment with ascorbate rescues mice from heat stroke-induced death by attenuating systemic inflammatory response and hypothalamic neuronal damage. Free Radic Biol Med. 2016;93:84–93. doi:10.1016/j.freeradbiomed.2015.12.017

7. T.-Y. Kao; C.-C. Chio; M.-T. Lin, DDS. Hypothalamic Dopamine Release and Local Cerebral Blood Flow During Onset of Heatstroke in Rats. Stroke. 1994;25(12):248386. doi:10.1161/01.STR.25.12.2483

8. Buggy DJ, Crossley AWA. Thermoregulation, mild perioperative hypothermia and post-anaesthetic shivering. Br J Anaesth. 2000;84(5):615–628. doi:10.1093/bja/84.5.615

9. Wright HE, Selkirk GA, McLellan TM. HPA and SAS responses to increasing core temperature during uncompensable exertional heat stress in trained and untrained males. Eur J Appl Physiol. 2010;108(5):987–997. doi:10.1007/s00421-009-1294-0

10. Barrett, Kim E., Susan M. Burman Hlb and JY. Ganog’s Review of Medical Physiology. 26th ed. Mc Graw Hill education, LANGE; 2022.

11. Stewart S, Keates AK, Redfern A, McMurray JJV. Seasonal variations in cardiovascular disease. Nat Rev Cardiol. 2017;14(11):654–664. doi:10.1038/nrcardio.2017.76

12. Iba, T., Helms, J., Levi M et al. Inflammation, coagulation, and cellular injury in heat-induced shock. Inflamm Res. 2023;72:463–473.

13. Proctor EA, Dineen SM, Van Nostrand SC, et al. Coagulopathy signature precedes and predicts severity of end-organ heat stroke pathology in a mouse model. J Thromb Haemost. 2020;18(8):1900–1910. doi:10.1111/jth.14875

14. Hifumi T, Kondo Y, Shimazaki J, Oda Y, Shiraishi S, Wakasugi M. Prognostic significance of disseminated intravascular coagulation in patients with heat stroke in a nationwide registry. 2024;44(2018):306–311. doi:10.1016/j.jcrc.2017.12.003

